# Reactogenicity of COVID-19 vaccine in hemodialysis patients: a single-center retrospective study

**DOI:** 10.1101/2021.11.21.21266561

**Authors:** Gaetano Alfano, Niccolò Morisi, Francesco Fontana, Roberta Scarmignan, Laura Tonelli, Camilla Ferri, Martina Montani, Silvia Giovanella, Giulia Ligabue, Giacomo Mori, Erica Franceschini, Giovanni Guaraldi, Gianni Cappelli, Riccardo Magistroni, Gabriele Donati

## Abstract

**Introduction:** Some hemodialysis patients are reluctant to COVID-19 for the development of adverse events (AEs). The aim of this study was to verify the safety of mRNA-1273 vaccine in hemodialysis patients.

**Methods:** We conducted a retrospective analysis of in-center hemodialysis patients who underwent mRNA-1273 vaccine from March 1^st^ to April 30^th^, 2021. All AEs occurring after the first and the second doses were collected and classified as local or systemic.

**Results:** Overall, 126 patients on chronic maintenance dialysis without a prior COVID-19 diagnosis were vaccinated with two doses of mRNA-1273 vaccine. Mean age was 68 (IQR, 54,7-76) years and 53.6% of patients were aged ≥ 65 years. During the observational period of 68 (IQR, 66-70) days, AEs occurred in 57.9% and 61.9% of patients after the first dose and second dose, respectively. The most common AEs were: injection-site pain (61.9%), erythema (4.8%), itching (4.8%), swelling (16.7%), axillary swelling/tenderness (2.4%), fever (17.5%) headache (7.9%), fatigue (23.8%), myalgia (17.5%), arthralgia (12.7%), dyspnoea (2.4%), nausea/vomiting (7.1%), diarrhoea (5.6%), shivers (4%) and vertigo (1.6%).

The rates of local AEs were similar after the first and second doses (P=0.8), whereas systemic AEs occurred more frequently after the second dose (P=0.001). Fever (P=0.03), fatigue (P=0.02) and nausea/vomiting (P=0.03) were significantly more frequent after the second dose of the vaccine. There were no age-related differences in the rate of AEs. Overall, vaccine-related AEs in hemodialysis patients seem to be lower than in the general population.

**Conclusion:** RNA-1273 vaccine was associated with the development of transient AEs after the first (57.9%) and second doses (61.9%) in patients on chronic maintenance hemodialysis. Systemic AEs were more common after the second dose. Overall, all AEs lasted for a few days, without any apparent sequelae.

## Introduction

Coronavirus Disease-2019 (COVID-19) is a novel infectious disease that carries a high burden of mortality and morbidity worldwide^1^. In the absence of a specific treatment for COVID-19, vaccination is currently the most effective strategy to prevent this disease^2^. Highly effective vaccines against SARS-CoV-2 infection have been developed and administered globally for a full-vaccine coverage strategy. To attain the goal of reducing COVID-19 related mortality, current policies prioritized vaccination of frail populations including patients on maintenance hemodialysis^3^. This group of patients is particularly at risk of COVID-19 because includes subjects with advanced age and compromised immunity^4^, moreover, the thrice-a-week in-center haemodialytic treatment carries a high risk of cluster infection^5,6^. For these reasons, the immunization of hemodialysis patients has been recognized as a public health priority worldwide and a mass vaccination with RNA-platform such as mRNA-1273 vaccine (previously known as vaccine Moderna) has been soon conducted after the release of regulatory agencies’ authorization. Safety and efficacy of mRNA-1273 vaccine has been tested in phase III clinical trial (ClinicalTrials.gov n. NCT04470427) recruiting 30.351 participants. The promising results showed 94.1% efficacy at preventing Covid-19 illness, including severe disease^7,8^.

Vaccine-related adverse events (AEs) were usually mild or moderate in intensity and resolved within a few days^7^. The most commonly AEs were injection-site pain (92%), fatigue (70%), headache (64.7%), myalgia (61.5%), arthralgia (46.4%), shivers (45.4%), nausea/vomiting (23%), axillary swelling/tenderness (19.8%), fever (15.5%), swelling (14.7%) and erythema (10%). Nevertheless, the fear of developing vaccine-related symptoms led patients to deny vaccination or additional booster doses. A recent nationwide vaccine acceptability survey conducted in 150 facilities in the United States reported that about half of patients who were vaccine-hesitant expressed concerns about vaccine-side effects^9^. This perception is contradicted by recent vaccine safety data reporting the incidence of severe AEs was similar in mRNA-1273 (1.0%, 147 events) and placebo (1.0%, 153 events) groups during the study period^10^. However, no studies have been conducted to evaluate the reactogenicity of the mRNA-1273 vaccine in patients on maintenance dialysis. To inform public health and clinical practice, we investigated the safety of the mRNA-1273 vaccine in a cohort of hemodialysis patients.

## Methods

Study population included patients aged ≥18 years on chronic maintenance hemodialysis at the University Hospital of Modena, Italy. Anti-SARS-CoV-2 vaccination was performed in all patients without a prior COVID-19 diagnosis and without signs of ongoing infection who provided written consent from March 24^th^ to April 30^th^, 2021.

All charts of these patients were reviewed retrospectively.

This study has been authorized by the local Ethical Committee of Emilia Romagna (n. 839/2020).

RNA-1273 vaccine contains a molecule of messenger RNA (mRNA) carrying instructions encoding the spike protein, a protein on the surface of the SARS-CoV-2 virus which the virus needs to enter the body’s cells. Once the human cells have produced the spike protein, the immune system recognizes this protein as foreign and produces antibodies and activates T cells to attack it.

This vaccine was found to be safe and efficacious (94.1%) in preventing symptomatic, laboratory-confirmed COVID-19 in a large, randomized-controlled trial^7^ and subsequent observational studies^11,12^. To facilitate its availability and its use, Food and Drug Administration (FDA) and European Medicine Agency issued an emergency use authorization in the United States (December 2020) and Europe (January 2020), respectively^10,13^.

In our patients RNA-1273 vaccine was administered as two intramuscular injections, 28 days apart, by dialysis nurses. Injection of the vaccines was provided 30 minutes before the start of the dialysis session on the non-arteriovenous fistula arm. To optimize the number of vaccine doses, some patients were vaccinated at the end of the dialysis session if no hemodialysis-associated complications (hypotension, nausea, vomiting) occurred during the treatment. Patients were monitored on-site for at least 15 minutes after the vaccine injection. Subjects who had severe allergic reactions or any type of immediate allergic reaction to drugs were monitored for at least 30 minutes. Anticoagulant therapy was left unchanged when vaccination was performed before the dialysis session.

### Adverse events

AEs were considered related to the vaccine when they occurred soon after the vaccination. The duration of the side effects was counted as a whole 24 hour period. Fever was classified as a symptom only when body temperature was above 37.5 °C.

#### Statistical analysis

Baseline characteristics were described using median and interquartile range (IQR) or mean and standard deviation (SD). The percentage was used to describe categorical variables. The Student’s t-test and chi-square or Fisher’s test were used to compare continuous compare and categorical variables between groups, respectively.

A P value of <0.05 was considered statistically significant. All statistical analyses were performed using SPSS 24® statistical software.

## Results

RNA-1273 vaccine was administered to 126 patients on chronic maintenance hemodialysis. Baseline characteristics of participants are shown in Table 1. Median age was 68 (IQR, 54.7-76) years and more than half of patients (53.6%) were aged ≥65 years. Seventy-one patients were male (53.6%) and the majority of the patients were of Caucasian origin (87.3%). Median observation time from the second dose of the vaccine to the end of the study was 68 (IQR, 66-70) days.

**Table 1.** Demographic and clinical characteristics of hemodialysis patients who underwent RNA-1273 vaccine

| **Basal characteristics** | **All patients**<br>**n =126** |
| --- | --- |
| Age (yr) | 68 (54.7-6) |
| (range) | 19-92 |
| ≥ 65 yr | 71 (56.3) |
| Males, n. (%) | 71 (56.31) |
| Ethnic origin, n. (%) |  |
| Caucasian | 110 (87.3) |
| African | 15 (11.9) |
| Hispanic | 1 (0.8) |
| Etiology of ESRD, n. (%) |  |
| Nephrosclerosis | 54 (42.9) |
| Glomerulonephritis | 26 (20.6) |
| Diabetes | 14 (11.1) |
| ADPKD | 4 (3.2) |
| Nephrotoxic | 4 (3.2) |
| Pyelonephritis | 4 (3.2) |
| Interstitial | 3 (2.4) |
| HIVAN | 2 (1.6) |
| Others | 10 (7.9) |
| NA | 5 (4) |
| HD treatment schedule, n. (%) |  |
| 3 times per week | 115 (91.2) |
| 2 times per week | 7 (5.5) |
| 4 times per week | 4 (3.1) |
| Infectious disease, n. (%) |  |
| HBV | 3 (2.3) |
| HCV | 3 (2.3) |
| HIV | 2 (1.5) |
| Time elapsed from the first to the second dose of vaccine, day | 28 (28-28) |
| Follow-up, day | 68 (66-70) |

ESRD denotes end-stage renal disease; HBV, hepatitis B virus; HCV, hepatitis C virus

### First dose

After the first dose of RNA-1273 vaccine, local AEs occurred in 68 (53,9%) hemodialysis patients whereas systemic AEs in only 20 of them (15.8%).

As shown in Figure 1A, the most common local symptoms were injection-site pain (50.7%) followed by local swelling (9.5%), erythema (3.1%) and itching (3.1%).

**Figure 1.**
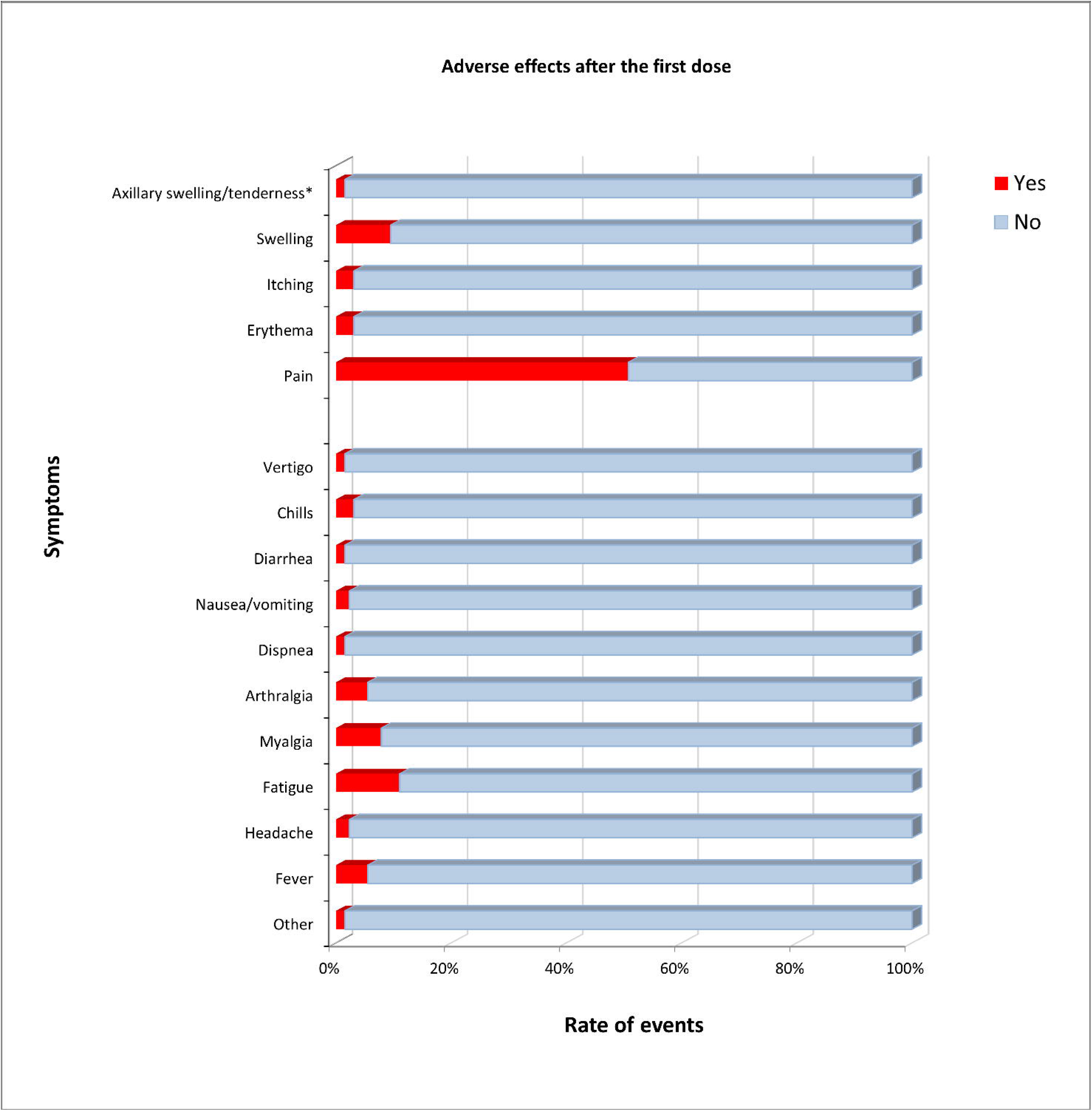
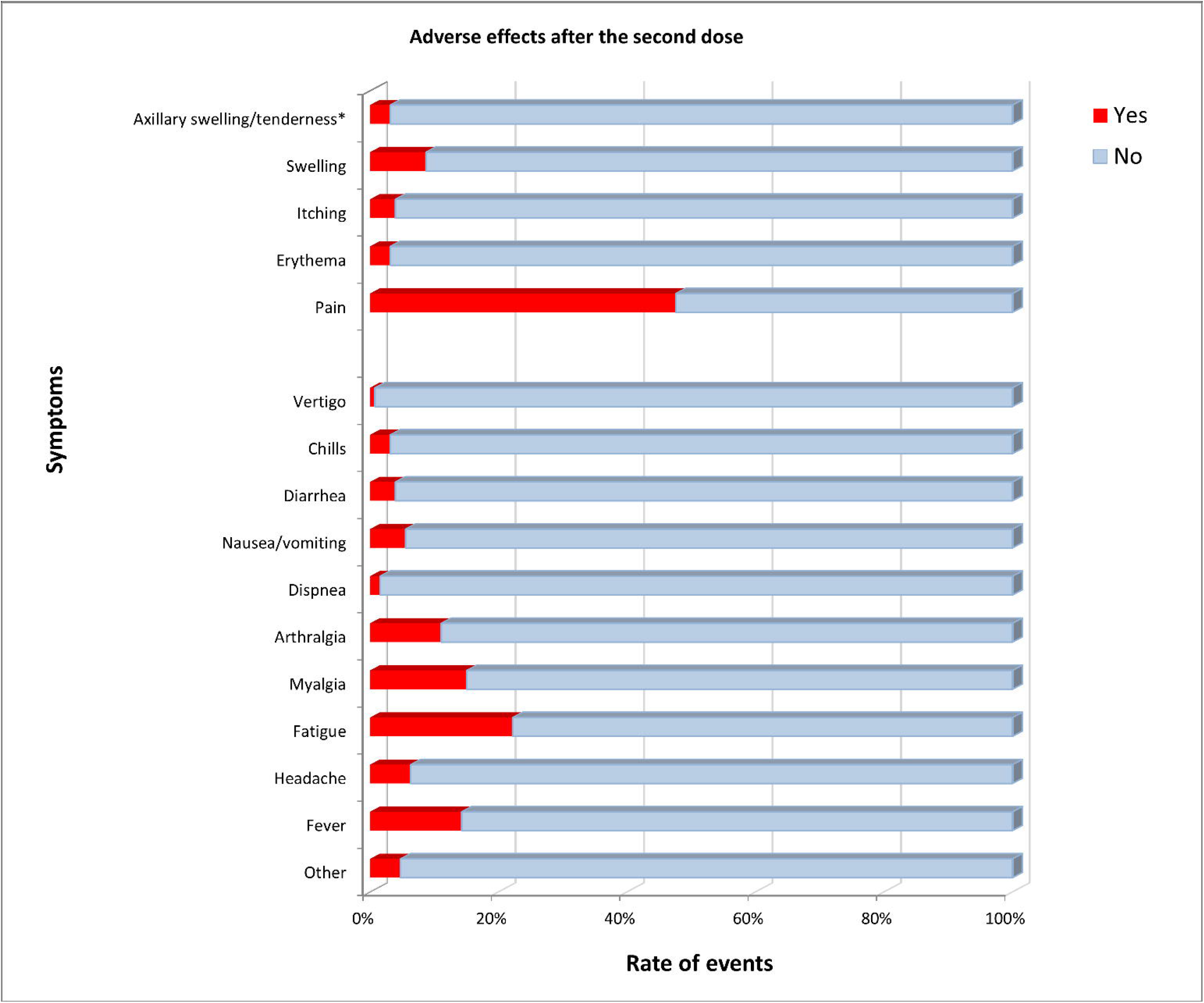
Rate of AEs after the first (**A**) and second dose (**B**) of mRNA-1273 vaccine

Fatigue (11.1%), myalgia (7.9%) fever (5.1%) and arthralgia (5.1%) were the most common systemic AEs experienced by hemodialysis patients.

### Second dose

The second dose was associated with local (51.6%) and systemic (34.1%) symptoms. The most common local AEs were site injection-site pain (47.6%) followed by local swelling (8.7%) and itching (3.9%). Systemic AEs included fatigue (22.2%), fever (14.2%) and myalgia (15.07%) (Figure 1B).

### Difference between first and second dose

Time elapsed from the first dose and second dose was 28 (IQR, 28-28) days. No differences were found in terms of local AEs between the first and second doses of vaccine (P=0.8), whereas the second dose was significantly associated with more systemic AEs than the first dose. (P=0.001).

As detailed in Table 2 and Figure 2, fever (P=0.033), fatigue (P=0.02) and nausea/vomiting (P=0.03) occurred more frequently after the second dose of vaccine.

**Table 2.** Rate of AEs after the first and second dose

|  | First dose |  | Second dose |  | p value |
| --- | --- | --- | --- | --- | --- |
| Any local AR. n. (%) | 68 | (53.9) | 65 | (51.5) | 0.80 |
| Pain. n. (%) | 64 | (50.7) | 60 | (47.6) | 0.70 |
| Erythema. n. (%) | 4 | (3.1) | 4 | (3.1) | >0.99 |
| Itching. n. (%) | 4 | (3.1) | 5 | (3.9) | >0.99 |
| Swelling. n. (%) | 12 | (9.5) | 11 | (8.7) | >0.99 |
| Axillary swelling/tenderness*. n. (%) | 2 | (1.5) | 4 | (3.1) | >0.99 |
| Any systemic AR. n. (%) | 20 | (15.8) | 43 | (34.1) | **0.001** |
| Fever. n. (%) | 7 | (5.1) | 18 | (14.2) | **0.03** |
| Headache. n. (%) | 3 | (2.3) | 8 | (6.3) | 0.21 |
| Fatigue. n. (%) | 14 | (11.1) | 28 | (22.2) | **0.02** |
| Myalgia. n. (%) | 10 | (7.9) | 19 | (15) | 0.11 |
| Arthralgia. n. (%) | 7 | (5.1) | 14 | (11.1) | 0.17 |
| Dispnea. n. (%) | 2 | (1.5) | 2 | (1.5) | >0.99 |
| Nausea/vomiting. n. (%) | 3 | (2.3) | 7 | (5.5) | **0.03** |
| Diarrhea. n. (%) | 2 | (1.5) | 5 | (3.9) | 0.44 |
| Chills. n. (%) | 4 | (3.1) | 4 | (3.1) | >0.99 |
| Vertigo. n. (%) | 2 | (1.5) | 1 | (0.7) | >0.99 |
| Other | 2 | (1.5) | 6 | (4.7) | 0.28 |

\*Localized axillary swelling or tenderness ipsilateral to the vaccination arm

**Figure 2.**
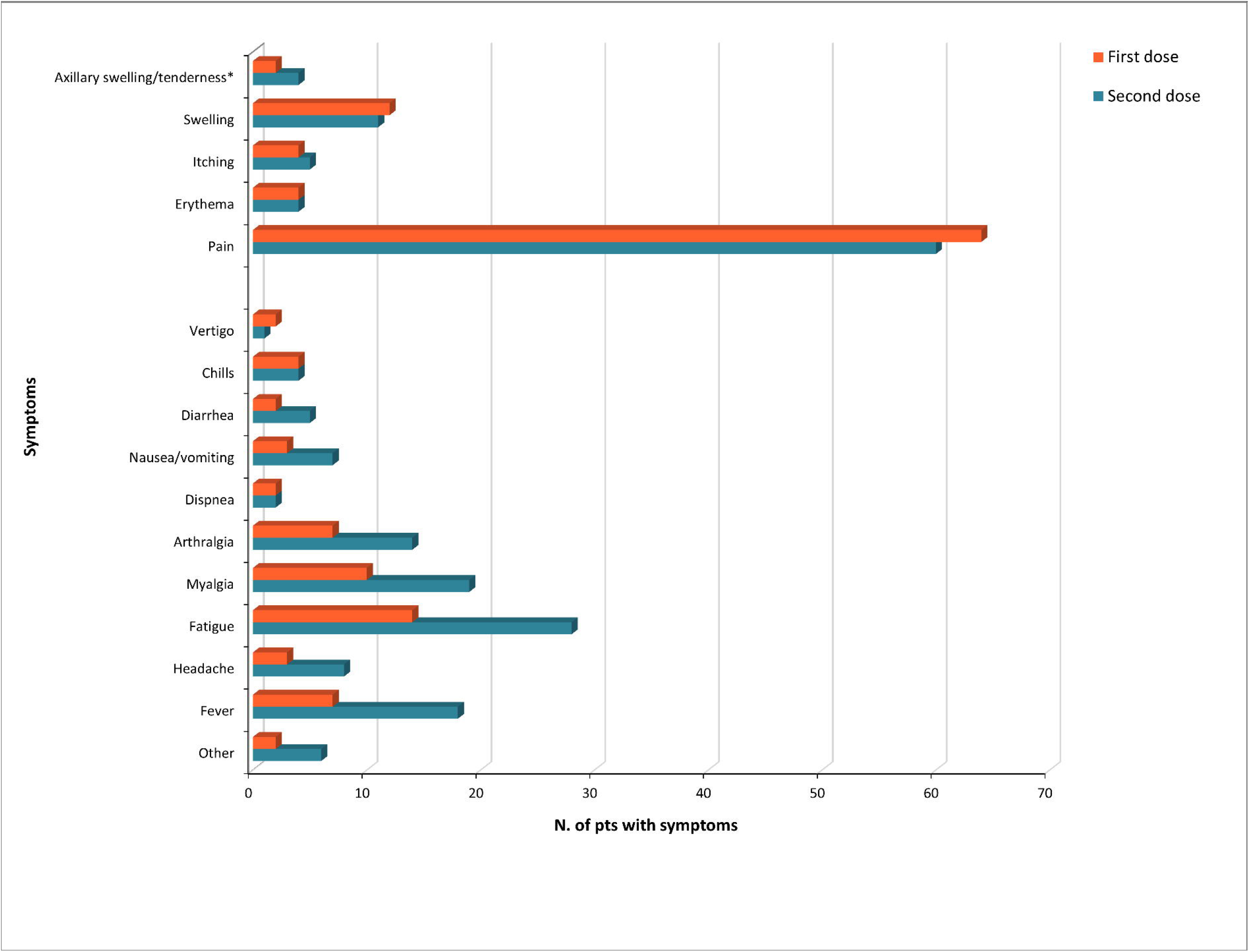
Patients who experienced AEs after the first and second dose

Seventy-three (57.9%) patients reported at least one AE after the first dose and 78 (61.9%) at least one AE after the second dose.

Analysis of the data detected statistically significant differences in duration of symptoms between the first and second doses regarding axillary swelling/tenderness (P=0.07) and diarrhea (P= 0.02). In both cases, these symptoms lasted longer after the second dose of the vaccine (Table 3). No differences in terms of symptoms were found between younger participants (18 to <65 years of age) and older participants (≥65 years of age) (Figure 3 A e B).

**Table 3.** Duration of AEs after the first and second doses

|  | Duration of AEs in days |  |  |  | p value |
| --- | --- | --- | --- | --- | --- |
|  | First dose<br>*mean (±SD)* |  | Second dose<br>*mean (±SD)* |  |  |
| Pain | 1.96 | (1.02) | 2.16 | (1) | 0.28 |
| Erythema | 1.75 | (0.95) | 3 | (1.4) | 0.2 |
| Itching | 2 | (0.81) | 2 | (0.7) | 1 |
| Swelling | 1.75 | (0.9) | 2.27 | (1.3) | 0.26 |
| Axillary swelling/tenderness* | 1 | (0) | 3.3 | (1.2) | **0.07** |
| Fever | 1.14 | (0.37) | 1.22 | (0.4) | 0.65 |
| Headache | 1.33 | (0.57) | 2 | (0.5) | 0.10 |
| Fatigue | 1.85 | (0.86) | 2.03 | (1) | 0.58 |
| Myalgia | 1.8 | (0.78) | 2.21 | (1) | 0.24 |
| Arthralgia | 2 | (0.8) | 2.42 | (1.1) | 0.32 |
| Dispnea | 2 | (1.4) | 2.5 | (2.1) | 0.81 |
| Nausea/vomiting | 1.33 | (0.57) | 1.57 | (0.8) | 0.61 |
| Diarrhea | 1 | (0) | 2.2 | (0.4) | **0.016** |
| Chills | 1.75 | (0.5) | 2.5 | (1) | 0.22 |
| Vertigo | 1.5 | (0.7) | 3 |  | 0.33 |

\*Localized axillary swelling or tenderness ipsilateral to the vaccination arm.

**Figure 3.**
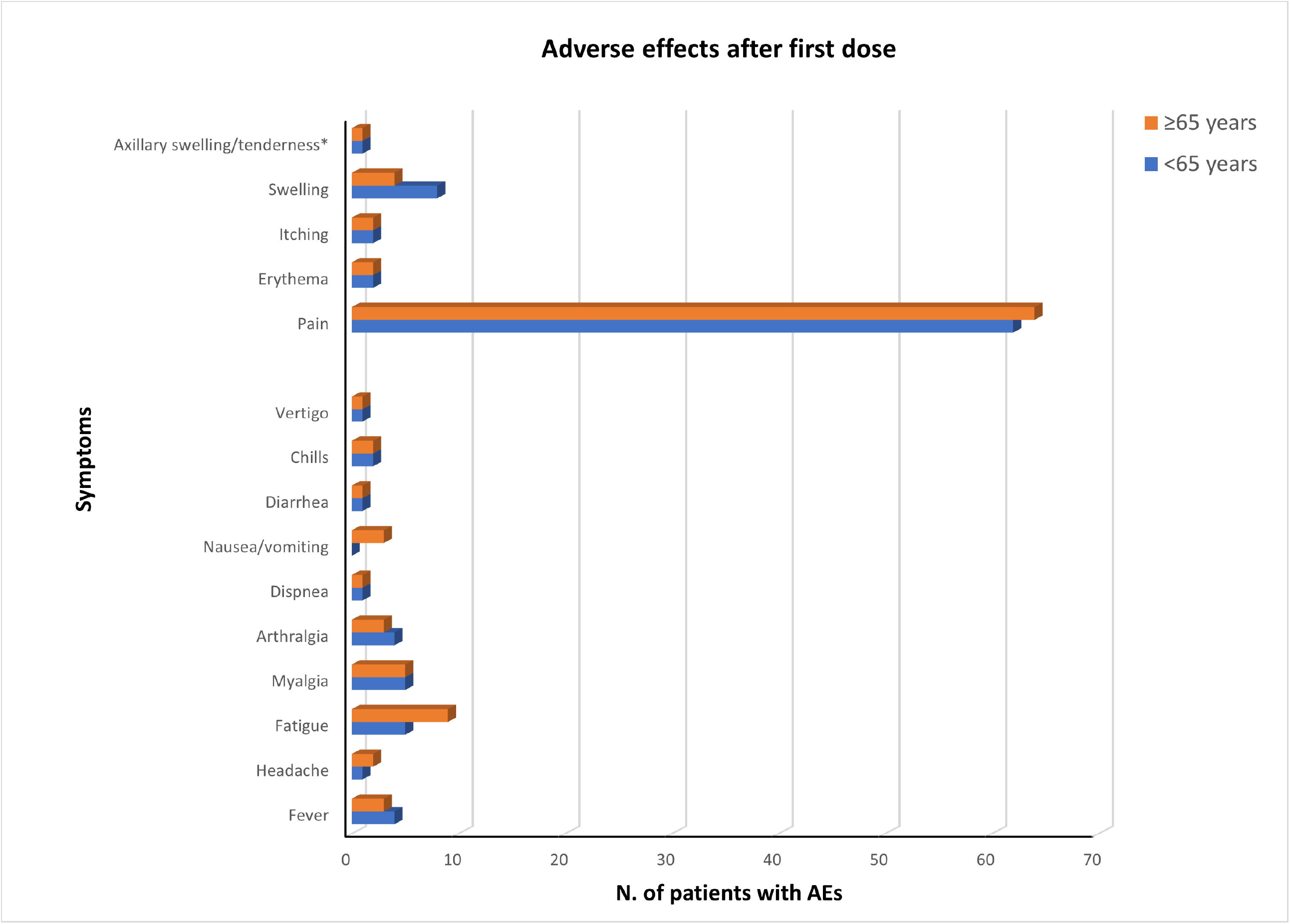
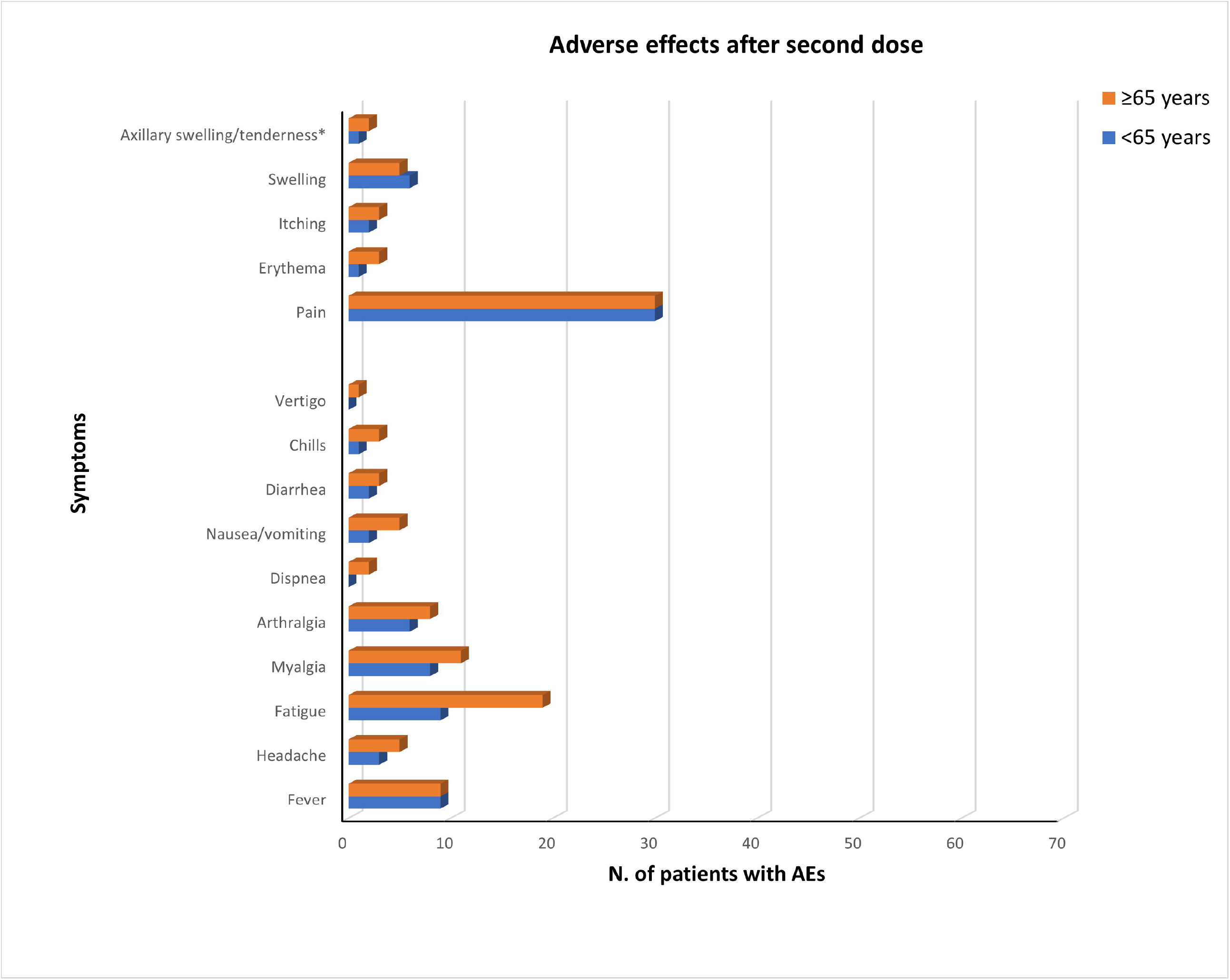
Patients stratified according to age who experienced AEs after the first (**A**) and second dose (**B**)

### Completed vaccination cycle

Overall, 71.4% of patients experienced vaccine-related AEs. In particular, the most commonly reported AEs were: injection-site pain (61.9%), erythema (4.8%), itching (4.8%), swelling (16.7%), axillary swelling/tenderness (2.4%), fever (17.5%) headache (7.9%), fatigue (23.8%), myalgia (17.5%), arthralgia (12.7%), dyspnoea (2.4%); nausea/ vomiting (7.1%), diarrhoea (5.6%), shivers (4%) and vertigo (1.6%).

Management of AEs occurred in outpatient settings without the need for hospitalization. Comparison of these findings with the data of phase III randomized trial^7,8^ showed that hemodialysis patients reported a lower rate of AE compared to the general population after the first dose (57.9% vs. 84.2%) and the second dose (61.9% vs. 88.6%).

Only one patient had herpes zoster reactivation after one week from the first dose. It healed after the administration of specific antiviral therapy.

## Discussion

mRNA-1273 vaccine is an effective tool of our armamentarium to prevent the severe consequences of COVID-19. This type of vaccine, based on RNA platform technology, has shown short- and long-term safety and efficacy in preventing symptomatic, laboratory-confirmed COVID-19 in a phase III trial^7,8^ and post-marketing studies^11,12^. Similar to other vaccines, RNA-1273 is associated with mild-moderate local and systemic AEs. Generally, these side effects developed within a few days after vaccination as a sign of an effective immune response against the foreign protein.

At present, the reactogenicity of mRNA-1273 of vaccine is unknown in the hemodialysis population, given their exclusion from the pre-marketing clinical trial. The low response rate to the vaccine and the high rate of COVID-19 infection after vaccination in the hemodialysis population,^17,18^ supports the hypothesis that the rate of AEs might be even lower than the general population because this cohort of patients is believed to have defects in humoral and cellular immunity.

In our study, we found that the mRNA-1273 vaccine was associated with local and systemic AEs in patients on in-center maintenance hemodialysis. About three-quarters (71.4%) of our patients experienced any type of AEs after the completion of the dual-dose SARS-CoV-2 vaccination. The rate of local reactions was higher than systemic AEs for both doses, without any statistically significant differences between the first and the second doses.

Injection-site pain, fatigue and myalgia were the most commonly reported side effects reported by our patients. Overall, these symptoms were transient and recovery occurred without sequelae, on average, within 2 days from the injection. As expected, the immune system of our patients, sensitized to the foreign protein, had a more robust response with the delivery of the second dose. A higher rate of AEs including fever (14.2% vs 5.1%), fatigue (22.2% vs 11.1%), myalgia (11.1% vs 22.2%), arthralgia (11.1% vs 5.1%) and headache (6.3%vs 2.3%) was indeed observed after the administration of the second.

We also investigate the age-related reactogenicity of RNA-1273 in our patients, because this vaccine showed a slightly lower frequency injection-site and systemic AEs in older participants (≥65 years of age) compared to younger participants (18 to <65 years of age). Our findings showed the rate and the duration of AEs were similar between the younger (18 to 64 years) and older (65– 84 years) hemodialysis patients. The reasons why the RNA-1273 vaccine elicited no age-related differences in the development of AEs are unclear. Likely, the burden of immunodepression induced by ESRD heavily affects the immune system of hemodialysis patients, and minimizes the differences due to aging.

Taking all together, mild-moderate vaccine-related AEs are frequent in hemodialysis patients and, consequently, healthcare workers in the dialysis units must learn to cope with them. Generally, the detection of severe systemic symptoms (i.e., fever, dyspnoea, nausea or vomiting) requires immediate attention because these symptoms are common to a large spectrum of conditions including infections, fluid overload, and electrolytes disturbance as well as COVID-19. Thus, we suggest a prudential behavior in hemodialysis patients presenting with fever and fatigue after vaccination, in order to prevent a potential COVID-19 outbreak in the dialysis unit.

Lastly, we noted a low rate of AEs of SARS-CoV-2 vaccine in hemodialysis patients after the first dose (57.9% vs. 84.2%) and second dose (61.9% vs. 88.6%) when compared to the general population. This low-level reactogenicity should be partially due to the burden of comorbidities, advanced age and suboptimal immune response of hemodialysis patients against SARS-CoV-2 antigen. In support of this theory, there is evidence that the effectiveness of hepatitis B^16^, influenza^17^ and SARS-CoV-2 vaccination^18,15^ is reduced in this frail population.

Some limitations of the study should be enunciated. The retrospective nature of the analysis and the small sample size could lead to unintended bias in the correct estimation of vaccine-related AEs.

Measure of AEs magnitude was not collected in our patients, albeit none required acute hospital in-patient care after the administration of vaccine. Lastly, our study provides key information for healthcare workers to plan active surveillance on hemodialysis patients.

## Conclusion

RNA-1273 vaccine was associated with the development of transient AEs after the first (57.9%) and second dose (61.9%) of vaccine in patients on chronic maintenance dialysis. Systemic AEs were more common after the second dose than the first dose of vaccine. The duration of these symptoms lasted for a few days, without any apparent consequences. These data confirm the safety of the RNA-1273 vaccine in hemodialysis patients and support the promotion of COVID-19 vaccination in hesitant patients.

## Data Availability

All data produced in the present work are contained in the manuscript

## Acknowledgments

Special thanks are due to the nephrologists and nurses who managed the vaccination of hemodialysis patients within our dialysis unit.

